# Structural network embedding governs peritumor and distant pathological brain activity in glioblastoma

**DOI:** 10.64898/2026.05.05.26352433

**Authors:** Mona LM Zimmermann, Marike R van Lingen, Eva Koderman, Sébastien CH Dam, Lucas C Breedt, Dorien A Maas, Niels Verburg, Philip C de Witt Hamer, Arjan Hillebrand, Linda Douw

**Affiliations:** Amsterdam UMC location Vrije Universiteit Amsterdam, Anatomy and Neurosciences, Amsterdam, The Netherlands; Amsterdam UMC location Vrije Universiteit Amsterdam, Amsterdam Neuroscience, Amsterdam, The Netherlands; Cancer Center Amsterdam, Amsterdam, The Netherlands; Amsterdam UMC location Vrije Universiteit Amsterdam, Neurosurgery, Amsterdam, The Netherlands; Amsterdam UMC location Vrije Universiteit Amsterdam, Clinical Neurophysiology and MEG Center, Amsterdam, The Netherlands

**Keywords:** cancer neuroscience, structural connectivity, brain activity, neuron-glioma interactions, tumor-brain cross-talk, connectome, brain network

## Abstract

Glioblastomas integrate into the brain globally, where they provoke neuronal hyperactivity to enhance tumor growth and invasion. Communication of glioblastomas with neurons is not only present locally, but has preclinically been shown to extend towards the contralateral hemisphere through white matter tracts. However, it remains unknown how the distant hyperactivity that is often found in patients relates to structural embedding of the tumor into the larger brain network.

29 newly diagnosed IDH-wildtype glioblastoma patients and 25 age and sex matched healthy controls were included. To define structural tumor embedding, we overlayed each patient-specific tumor mask with a normative structural connectome obtained from diffusion MRI. We identified the average number of streamlines intersecting the tumor, extracting the tumor’s average tract density (‘Lesion-Tract Density Index’, L-TDI). For a subgroup of patients (*n* = 17), we determined structural embedding directly from diffusion scans and subsequent tractography. To identify regions connecting to the tumor, we seeded from each patient’s tumor rim outside FLAIR hyperintensities in the white matter to the 210 cortical regions of the Brainnetome atlas. We then counted the number of tumor-connecting regions, termed PATNET hereafter. Finally, participants underwent eyes-closed resting-state magnetoencephalography. We used broadband power as a proxy for neuronal spiking activity of each cortical region. To capture deviant brain activity in tumor and non-tumor regions, we regionally standardized broadband values using controls. We then sought to establish an association of deviant peritumor activity with both L-TDI and PATNET. Subsequently, we investigated whether tumor-connected regions showed more deviant activity than unconnected regions. Finally, we explored the clinical relevance of L-TDI and PATNET.

Greater structural tumor embedding significantly related to more deviant peritumor activity (*rho_LTDI_*= 0.47, *P_LTDI_* = .010; *rho_PATNET_* = 0.54, *P_PATNET_* = .024), with larger tumors showing greater embedding and more hyperactivity than smaller tumors. Furthermore, distant tumor-connected regions showed more hyperactivity than unconnected regions, but only in patients with peritumor hyperactivity (*F*(1,15) = 11.02, *P* =.005). Finally, higher PATNET associated with lower KPS (*U* = 61.5, *P* = .015).

Glioblastomas’ structural embedding explains hyperactivity around the tumor and in distant cortical regions, such that distant hyperactivity occurs primarily when there is tumor hyperactivity and the region is structurally connected to the tumor. Moreover, patient-specific tumor embedding relates to functional status.

## Introduction

IDH-wildtype glioblastoma is the most aggressive primary brain cancer, causing a wide range of life-altering symptoms that remain poorly understood due to extensive tumor heterogeneity and a lack of reliable tumor-specific markers.^1–3^ A key open question is how a tumor’s structural embedding in the broader brain network relates to disturbances in peritumor and distant brain activity and connectivity, as these disturbances are known to affect patients’ functional status, cognitive performance, and quality of life.^4–6^

Glioblastomas directly integrate into the brain environment.^7–10^ Paracrine factors and direct neurogliomal synapses form a bidirectional channel of communication between the tumor and the brain, via which higher neuronal spiking activity enhances tumor proliferation and invasion, while the growing tumor also provokes neuronal activity. Peritumor hyperactivity (i.e. hyperactivity measured in and directly around the tumor) associated with this communication can be indirectly and non-invasively measured in patients using magnetoencephalography (MEG). We have shown that (peri)tumor brain activity as operationalized through broadband power of the MEG signal is higher than normal, relates to paracrine factors excreted in neuron-glioma interactions, and significantly associates with shorter survival.^11–14^ Others have used different proxies of brain activity based on MEG signals, finding similar patterns.^15,16^

Importantly, the interactions of the tumor with the brain go far beyond the tumor-surrounding tissue: even neurons in the contralateral hemisphere innervate the tumor in animal models.^17,18^ In turn, tumor connectivity to distant regions may impact their neuronal activity as well.^17^ In some patients, we indeed observed disturbances in brain activity and connectivity even in distant regions.^9,11,13^ These disturbances include hyperactivity and functional network deviations, which are relevant for clinical and cognitive performance.^13,19,20^ However, it remains unknown what drives these distant disturbances, and why not all patients have them.

Potentially, the extent of tumor embedding into the structural brain network explains some of these heterogeneous hyperactivities. Glioblastomas occur more often in cortical regions that are intrinsically more functionally connected^19,21,22^, yet it is unclear whether this also holds for the structural connections that have been found relevant for glioblastoma in preclinical studies.^8–10^ It has become clear that structural tumor embedding, as operationalized by overlapping tumor masks with voxel-wise tractography data of healthy controls (the ‘Lesion -Tract Density Index’, L-TDI), is somewhat predictive of overall survival (OS).^17,18,23–25^ But do structural connections between the tumor and distant regions explain hyperactivity throughout the brain, and are these relevant for functional outcomes and survival?

Here, we investigated how structural tumor embedding based on normative as well as patient-specific tractograms relates to peritumor and distant brain hyperactivity. We hypothesized, firstly, that glioblastomas preferentially occur in regions with greater structural network embedding. Secondly, we expected that tumors’ structural network embedding governs pathologically high brain activity around the tumor and in distant tumor-connected, but not unconnected, regions. Finally, we hypothesized that greater network embedding associates with worse prognosis and clinical performance of patients.

## Materials and methods

### Participants

Patients were included from an ongoing observational study on diffuse glioma at Amsterdam UMC (see Table S1 for an overview of other publications on this growing cohort). Inclusion criteria were 1) age > 18 years, 2) no comorbidities of the CNS or neuropsychiatric disorders and 3) able to provide written informed consent. Selection criteria for the current analyses were 1) patients with confirmed isocitrate dehydrogenase (IDH), wildtype, unilateral glioblastoma^26^ and 2) availability of preoperative MEG recording on an ELEKTA system (Neuromag Oy, Helsinki, Finland). Healthy controls (HCs) were selected from a study that used the same MEG protocol^27^, and were matched to patients based on age and sex at the group-level. All participants gave written informed consent and the studies were approved by the VUmc Medical Ethical Committee and conducted according to the Declaration of Helsinki.

To correlate tumor occurrence with normative structural embedding, we additionally used the publicly available glioblastoma dataset from The Cancer Genome Atlas (TCGA) in combination with the Cancer Imaging Archive (tumor masks and patient characteristics available at GDC Data Portal Homepage, *n* = 102).^28,29^

### MRI acquisition

All patients were scanned before the start of any treatment on a 3T GE Discovery MR750 magnet (General Electric, Milwaukee, USA) with an 8-channel head coil. A high-resolution 3D T1-weighted sequence (FSPGR; TR = 4.6 ms; TI = 450 ms; TE = 2 ms; flip angle = 15°; 1 mm^3^ isotropic resolution; 174 sections) was acquired with and without gadolinium injection. A FLAIR image was also acquired (TR = 8000 ms; TI = 2340 ms; TE = 130 ms; flip angle = 90°; 1×1.2×1.2 mm resolution).

A subset of patients also underwent a single-shell diffusion MRI sequence in the same session (24 directions at b = 750 s/mm^2^; 4 b0 s/mm^2^; TR = 7.2 s; flip angle = 90°; 2 mm isotropic resolution; no gap).

### Tumor segmentation

The PICTURE toolbox, a deep learning algorithm,^30^ was used to segment each tumor based on T1-weighted scans with contrast, FLAIR images and optionally T2-weighted scans. The output is a mask consisting of three components: the necrotic core, enhancing tumor component, and FLAIR hyperintensity. We defined the tumor mask as the binarized union of the necrotic core and enhancing tumor component. This tumor mask was then overlapped with the cortical regions of the Brainnetome atlas^31^ in native MRI space (supplementary materials). Atlas regions were deemed tumor-invaded if at least 20% of their volume overlapped with the tumor mask.

Tumor masks for the TCGA dataset were available through the Brain Tumor Segmentation (BraTS) challenge.^28,29^ A tumor occurrence map was created in MNI space, where every voxel’s value reflects the number of patients with a tumor located there.^32,33^

### Normative Lesion Tract Density

We used the Lesion-Tract Density Index (L-TDI)^24,25^ as a measure of tumor embedding in the normative structural connectome. This normative tractogram was based on 985 healthy subjects and included more than 11.8 billion streamlines.^25^ We registered all patients’ tumor masks to MNI space, using FSL FLIRT, to overlay with the normative tractogram. Similar to previous studies^23–25^, we first created a whole-brain map of voxel-wise streamline weights by summing the number of streamlines passing through each voxel. We later used this map to investigate linkage between tumor occurrence and voxel-wise network embedding.

Next, we obtained the L-TDI in three steps.^24,25^ First, we identified the streamlines passing the tumor mask, using a publicly available script ran in Matlab (R2022b, Mathworks, Inc.).^34^ We then counted the number of tumor-intersecting streamlines that passed each voxel to quantify tumor-related tract density, thereby creating a lesion tract density map (Figure 1b). Finally, the mean of all non-zero voxels was calculated from this map.

**Figure 1.**
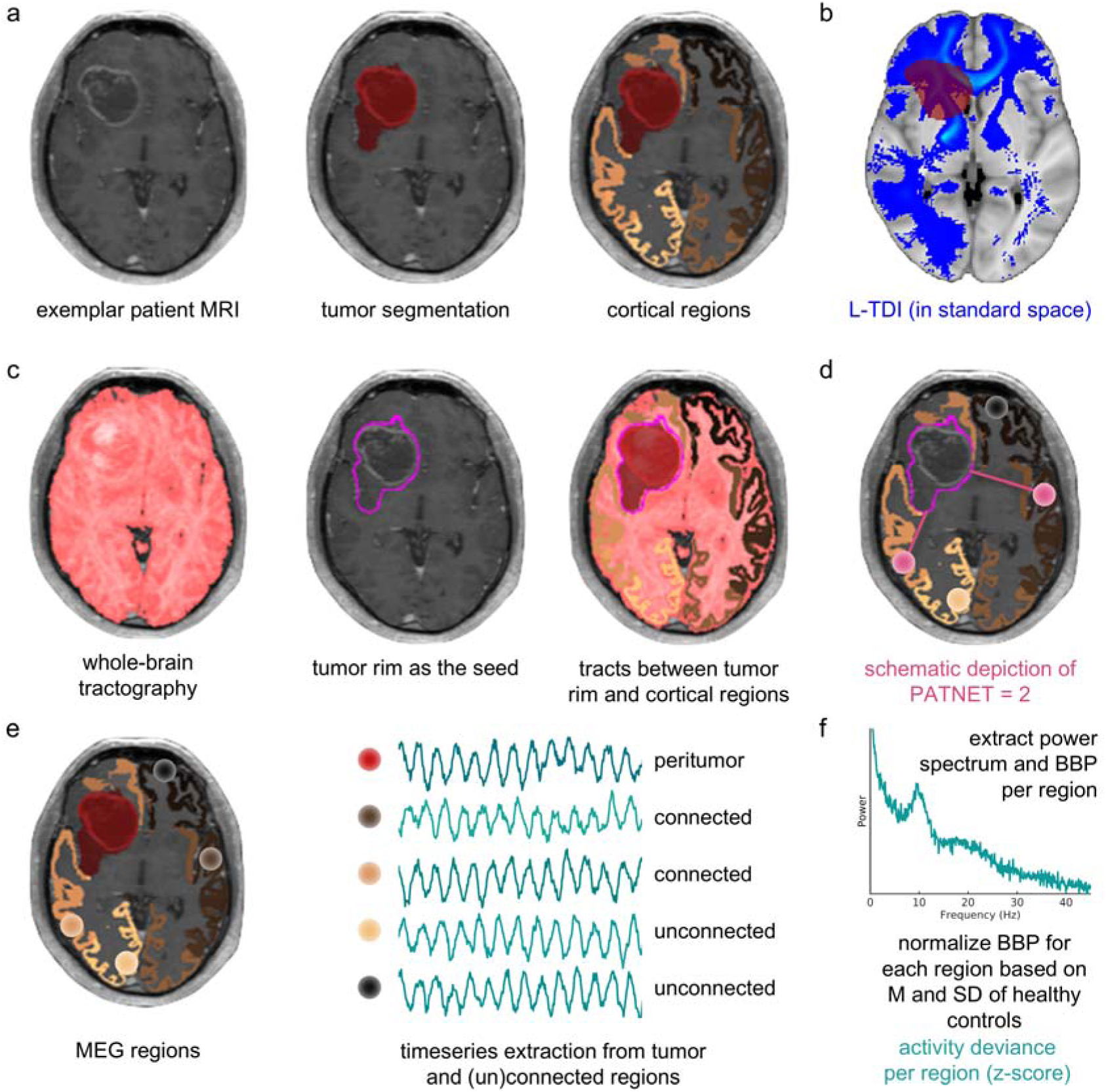
Schematic depiction of the analysis pipeline. (a) *Tumor mask.* Tumors were segmented automatically and overlaid onto Brainnetome atlas regions. Regions with ≥20% volumetric overlap with the tumor mask were classified as tumor-invaded. (b) *L-TDI.* Using a normative tractogram, we generated a tract density map per patient by counting streamlines passing through the tumor mask. L-TDI was then calculated as the mean density across all non-zero voxels, reflecting average tract density of tumor-intersecting tracts. (c,d) *PATNET.* From each patient’s own diffusion MRI tractogram, we extracted the tumor rim from a dilated tumor mask, retaining only white matter portions. We seeded tractography from these rim voxels to 210 atlas regions, defined tumor-connected regions as those receiving ≥1000 streamlines, and counted these as a measure of patient-specific tumor network embedding (PATNET). (e,f) *Brain activity.* We extracted timeseries from all 210 atlas regions and computed broadband power (0.5–48 Hz) using Welch’s method. Values were z-scored relative to healthy controls (M = mean; SD = standard deviation) to obtain a deviation measure. Peritumor activity was defined as mean broadband power across tumor-invaded regions.

### Patient-specific tractography

To extract patient-specific tumor embedding, we first applied the FreeSurfer recon-all-clinical pipeline (version 7.4.1),^35–38^ which uses deep learning to accurately reconstruct the cortical surface and the boundaries between gray and white matter, even in the context of a glioblastoma.

Diffusion MRI preprocessing steps included MP-PCA denoising, removing of Gibbs ringing artifacts, and eddy current distortion correction. We then used the Dhollander algorithm to estimate a tissue response function and Single-Shell 3-Tissue CSD (SS3T-CSD implemented in MRtrix3Tissue https://3Tissue.github.io) to determine the fiber orientation distribution (FOD) for each voxel in three tissue compartments (i.e. white matter, grey matter, and CSF). The resulting white matter FOD was subsequently intensity-normalized using mtnormalise. We used probabilistic tractography (IFOD2) to seed 100 million streamlines randomly within a whole-brain mask and performed spherical-deconvolution informed filtering of tractograms (SIFT2^39^) to improve the accuracy of the reconstructed streamlines and reduce false positives.

### Patient-specific tumor network embedding (PATNET)

We then sought to extract structural connectivity from the tumor to the 210 cortical atlas regions, to obtain patient-specific structural tumor network embedding (hereafter referred to as PATNET), and to allow linkage between structural embedding and brain activity. We first registered the tumor mask (encompassing the enhancing tumor and necrotic core, but now also the FLAIR hyperintensities to avoid artifacts in our streamline reconstruction) to FreeSurfer space. We then dilated the mask using fslmaths from FSL version 6.0.6.5, and subtracted the tumor itself from the resulting binary mask, leading to a peritumor rim. We multiplied this rim with the white matter mask in order to only seed from the white matter.

Finally, we extracted streamline probabilities between the tumor rim and each cortical region from the whole-brain tractogram (Figure 1c). We then classified tumor-connected regions as those with at least 1000 streamlines (from the 100 million seeds) between the tumor rim and the cortical region to reduce spurious connections, while excluding tumor-invaded cortical regions (i.e. regions that overlapped 20% or more with the tumor mask). To establish a single PATNET value per patient, we summed this number of tumor-connected regions (Figure 1d). Patients with PATNET values higher than the group median (42 tumor-connected regions) were considered to have strong PATNET.

### Magnetoencephalography

Resting-state, eyes-closed MEG data were recorded for five minutes using a 306-channel whole-head system (102 magnetometers and 204 gradiometers; Elekta Neuromag Oy, Helsinki, Finland) in a magnetically shielded room (Vacuumschmelze GmbH, Hanau, Germany). Signals were sampled at 1250 Hz, with online high-pass (0.1 Hz) and anti-aliasing (410 Hz) filters applied. Head position was checked using 4-5 head localization coils, and the scalp surface was obtained with a 3D digitizer (Fastrak, Polhemus, Colchester, VT) for coregistration with each participant’s anatomical MRI.

Initial preprocessing included offline application of cross-validation signal space separation^40^ and visual inspection of the raw data to remove malfunctioning or noisy channels, after which noise suppression was achieved using temporal signal space separation.^41^ Data were subsequently band-pass filtered between 0.5-48Hz. A single best-fitting sphere derived from the co-registered MRI was used as the volume conductor model for source reconstruction. Broadband activity was reconstructed using a scalar beamformer approach.^42,43^ The (normalized) beamformer weights were computed based on the lead fields, broadband data covariance, and a unity noise covariance matrix. Source time series were extracted for the centroids of the 210 cortical regions, resulting in one representative time series per region.^31^ Both patient and HC source-level data was then split into 22 epochs (total 288 sec) of 16384 samples each.

### Regional brain activity

Broadband power (BBP) served as a proxy for neuronal spiking activity, based on previous work on the link between neuronal spiking and BBP,^44^ as well as our earlier studies in glioblastoma patients.^11–13,45^ For every participant, we used Welch’s method with a hamming window and overlapping segments of 4096 samples to obtain the power spectrum over 0.5-48Hz per epoch. Power spectra were subsequently averaged over the 22 epochs to obtain one average spectrum per participant. Using the area under the curve of the power spectrum, we obtained the absolute BBP per brain region in each participant.

Patients’ regional BBP values were subsequently standardized based on the regional means and standard deviations of the HCs to obtain a measure of deviation (i.e. z-score, hereafter referred to as BBP_dev,_ Figure 1f). Patients with a mean BBP_dev_ > 1 in their tumor-invaded regions were considered to have peritumor hyperactivity, while patients with BBP_dev_ <= 1 were considered to have normal peritumor activity.

### Statistical analyses

All dataframe preparations and statistical analyses were performed using Python (3.12.3) and R studio (2024.12.1 Build 563) with supporting help of LLMs (ChatGPT and Claude).

First, we used Spearman’s rho to correlate the TCGA tumor occurrence map with normative streamline density across all voxels.

We also used Spearman’s rho to correlate mean tumor BBP_dev_ with L-TDI and PATNET across patients. We additionally tested the relationship between L-TDI and PATNET using Spearman’s rho.

Next, we averaged non-tumor BBP_dev_ over all tumor-connected regions per patient, as well as over unconnected regions. We then used type 3 mixed ANOVAs with averaged BBP_dev_ of the connected and unconnected regions as within-subject effects, and either peritumor activity (‘hyperactivity’ or ‘normal’) or PATNET (‘strong’ or ‘weak’) as between-subject effect.

Finally, we tested the clinical significance of L-TDI, PATNET and activity in tumor-connected regions for preoperative Karnofsky performance status (KPS), progression-free survival (PFS) and OS using independent t-tests (in case of violations of normality assumptions we used the Mann-Whitney U test, in case of unequal variance Welch’s t-test) and Cox-Proportional Hazards analyses, respectively. For the KPS we chose a cut-off at 90 to define groups, due to the spread of our data and based on clinical observation that this often distinguished between patients who do not experience any symptoms and patients with symptoms influencing daily life.

For all analyses, we evaluated the assumptions of the statistical tests and deemed results significant at *P* < .05.

## Results

### Participants

We included 29 patients with glioblastoma (of which 17 also underwent diffusion MRI), and 25 matched HCs (age *U* = 471.5, *P* = .059, sex (Χ*²*(1) = 1.56, *P* = .211). All participant characteristics can be found in Table 1. Characteristics of the TCGA patients can be found in Table S2.

**Table 1.** Participant characteristics.

| | Glioblastoma patients<br>($n = 29$) | Glioblastoma patients<br>PATNET subgroup<br>($n = 17$) | Matched healthy<br>controls<br>($n = 25$) |
| --- | --- | --- | --- |
| Sex (males/females) | 23/6 | 13/4 | 15/10 |
| Age in years (mean ± SD) | 59.3 ± 12.5 | 60.2 ± 11.4 | 51.7 ± 13.6 |
| Karnofsky Performance<br>Status (median (range)) | 100 (60 - 100) | 80 (70 - 100) | - |
| Epilepsy (yes/no) | 18/11 | 11/6 | - |
| Use of antiepileptic drugs<br>(yes/no) | 18/11 | 11/6 | - |
| Progression (yes/no) | 24/5 | 15/2 | - |
| Progression-free survival in<br>weeks (median (IQR)) | 42 (23 - 75) | 42 (20 - 61) | - |
| Tumor laterality (left/right) | 18/11 | 12/5 | - |
| Tumor volume corrected for<br>head size in mL (median<br>(range)) | 20.2 (2.6 - 82.4) | 24.3 (4.9 - 82.4) | - |

### Glioblastoma preferentially occurs in highly structurally embedded voxels

Using the tumor occurrence map from the TCGA cohort and the normative voxel-based tract density map, we established a strong and significant correlation between the frequency of glioblastoma occurrence per voxel and the number of streamlines running through that voxel (*rho*(69402310) = 0.72*, P* < .001, Figure 2). These findings indicate that glioblastoma preferentially occurs in highly structurally embedded locations.

**Figure 2.**
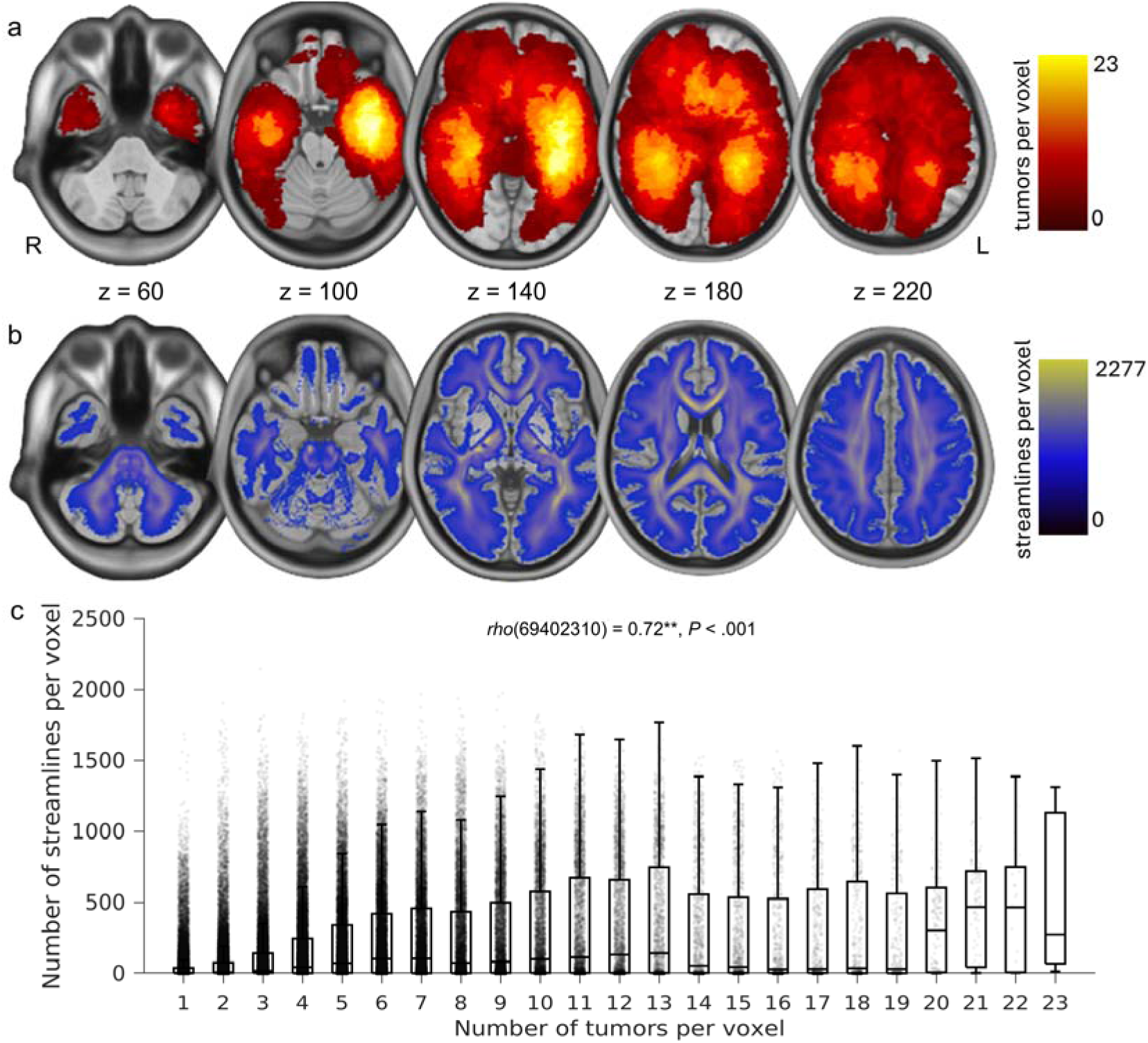
Association between tumor occurrence and number of streamlines. (a) *Tumor frequency.* Tumor frequency map of the TCGA cohort at different levels of the axial plane. Brighter colors indicate that a higher number of tumors occur in those voxels. (b) *Tract density map.* Normative tract density map indicating the number of streamlines passing every voxel at different levels of the axial plane. Brighter colors indicate that a higher number of streamlines pass those voxels. (c) Relationship between number of streamlines per voxel (y-axis) and tumor occurrence per voxel (x-axis) in the TCGA cohort. Each grey point represents one voxel.

### Higher network embedding governs deviant peritumor activity

We next sought to establish potential associations between peritumor hyperactivity and normative network embedding of the tumor (L-TDI) and the patient-specific network embedding of the tumor (PATNET). Indeed, L-TDI was significantly associated with peritumor activity deviance (*rho*(27) = 0.47, *P =* .010), as was PATNET (*rho*(15) = 0.54, *P* = .024, Figure 3a), such that greater structural embedding related to more hyperactivity around the tumor. Moreover, L-TDI and PATNET correlated strongly across patients (*rho*(44) = 0.76, *P* < .001), further suggesting minor differences between hypothesized tumor embedding based on normative data, and patient-specific embedding.

**Figure 3.**
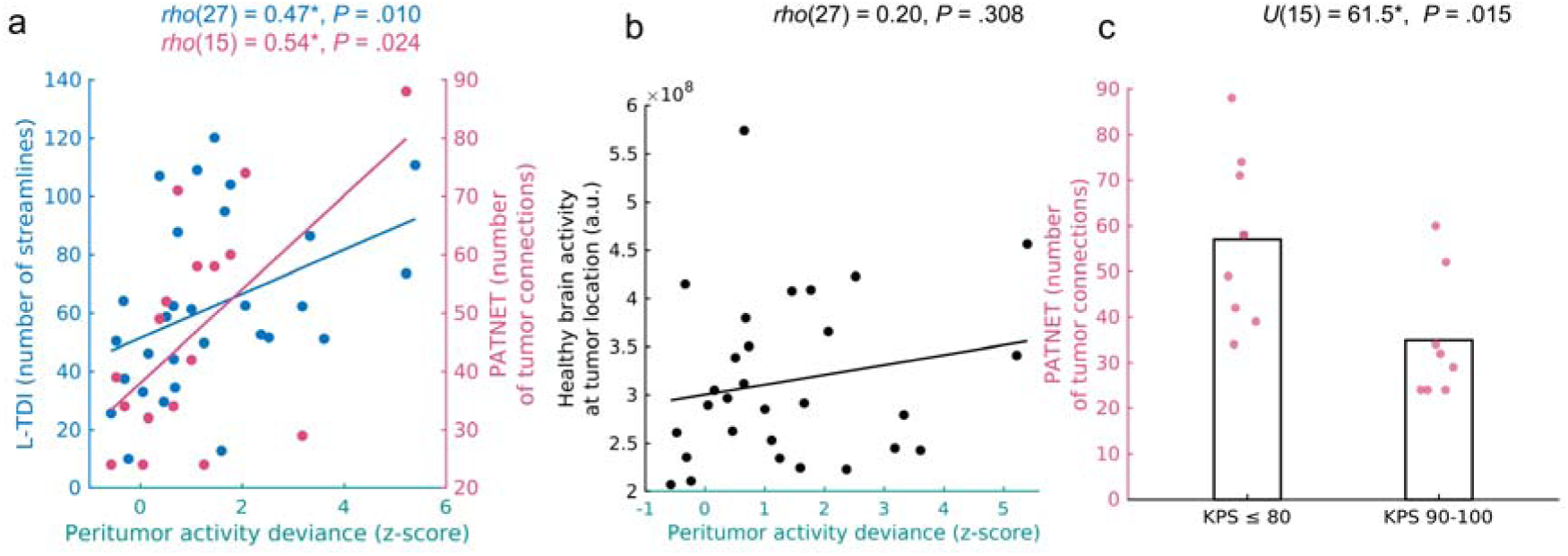
Tumor network embedding, peritumor activity deviance and functional status. (a) Correlation of L-TDI (blue, left y-axis) and PATNET (pink, right y-axis) with peritumor activity deviance. Every point represents a patient. (b) Correlation between regional brain activity in healthy controls and peritumor activity deviance. Every point represents a patient. (c) Comparison of PATNET values between low (<90) and normal KPS (90-100).

Of note, we also checked whether those patients with peritumor hyperactivity more often had their tumor in a location with normally high brain activity, by correlating average healthy brain activity at individual patients’ tumor locations with their peritumor activity deviance values. This revealed a non-significant correlation (*rho*(27) = 0.20*, P* = .308, Figure 3b), suggesting that this peritumor hyperactivity is not fully explained by intrinsic brain activity at the tumor location.

Subsequently, in a set of post-hoc tests, we investigated how tumor volume (corrected for head size by taking the MNI space tumor masks), age at diagnosis, sex and presence of epilepsy might relate to L-TDI, PATNET and peritumor hyperactivity. We found that tumor volume related positively to peritumor hyperactivity (*rho*(27) = 0.39, *P* = .039). PATNET (*rho*(15) = 0.52*, P* = .032) and L-TDI (*rho*(27) = 0.86, *P* < .001) also related to tumor volume, indicating unsurprisingly that larger tumors are more likely to intersect with more dense white matter pathways^25^. We also found age to negatively relate to PATNET (*rho*(15) = −0.48, *P* = .050), but not L-TDI (*rho*(27) = −0.08, *P* = .668), indicating that older individuals had less connected tumors, which was not due to their tumors occurring more often in regions that are normally less structurally embedded, potentially reflecting age-related variations in structural connectivity. Sex did not relate to L-TDI, PATNET or peritumor hyperactivity (Table S3).

We then explored the relevance of these brain features for patient outcomes. Presence of epilepsy did not relate to L-TDI, PATNET or peritumor hyperactivity (Table S3). We then tested whether L-TDI and PATNET were related to lower (<90) or high (90-100) KPS. While L-TDI did not relate to KPS (*t*(27) = 1.63, *P* = .115), strong PATNET related to lower KPS (*U*(15) = 61.5, *P* = .015, Figure 3c), suggesting that patient-specific tumor embedding might be more relevant for clinical functioning than tumor structural embedding based on normative data, although we did not test the significance of these differences directly.

In terms of survival, Cox proportional hazards analyses revealed non-significant hazards ratios concerning PFS and OS for both L-TDI and PATNET (Table S4).

### Tumor-connected regions show deviant brain activity only in patients with peritumor hyperactivity

We then tested the hypothesis that peritumor hyperactivity might relate to hyperactivity in distant but tumor-connected cortical regions. Firstly, we found a significant main effect of peritumor hyperactivity towards distant brain activity (*F*(1, 15) = 7.87, *P* = .013), such that patients with more peritumor hyperactivity also had higher brain activity in distant brain regions. There was also a significant main effect of connection to the tumor (*F*(1, 15) = 5.97, *P =* .027), indicating that tumor-connected regions generally showed higher brain activity. But most importantly, the significant interaction between peritumor hyperactivity and connection to the tumor (*F*(1, 15) = 11.02, *P* = .005, Table 2) confirms our hypothesis that distant brain activity is higher in tumor-connected regions than in unconnected regions, but only in patients who have peritumor hyperactivity (Figure 4a). To assess the specificity of this interaction to peritumor activity, we redid the analysis using a median split on tumor network embedding (PATNET, weak/strong) instead of peritumor hyperactivity, revealing similar but slightly weaker and more variable results (Figure 4b, Table 2). Taken together, these findings suggest that distant deviant hyperactivity occurs almost exclusively if (1) there is peritumor hyperactivity, and (2) the distant region is structurally connected to the tumor.

**Figure 4.**
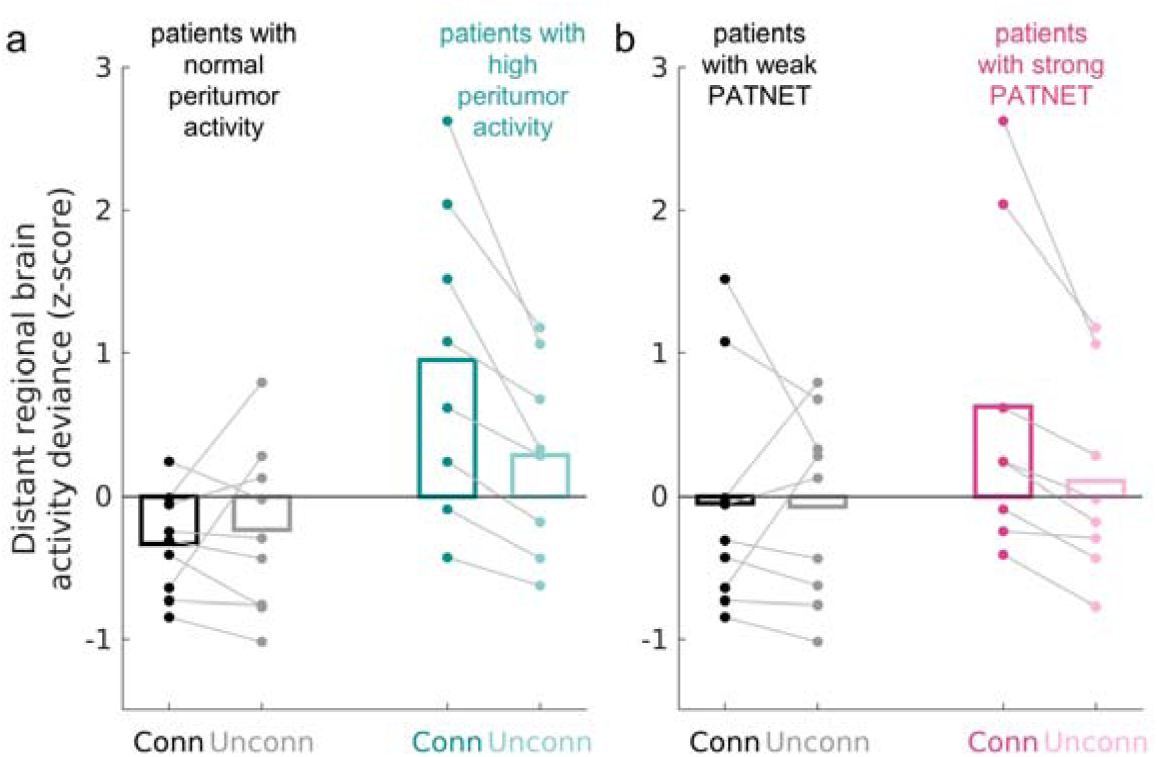
Interactions between tumor network embedding, peritumor activity deviance and distant regional brain activity deviance. (a) Comparison of activity deviance in tumor-connected regions (dark color) and unconnected regions (light color) for patients with peritumor hyperactivity (teal) and normal peritumor activity (gray). (b) Comparison of activity deviance in tumor-connected regions (dark color) and unconnected regions (light color) in patients with strong (pink) or weak (gray) PATNET. Lines between connected and unconnected bar plots indicate values of the same patient.

**Table 2.** Mixed ANOVAs on explaining activity in distant regions.

| Model 1: Peritumor activity | Effect | *df* | *F* | *P*-value |
| --- | --- | --- | --- | --- |
| **Between-subject** | Peritumor activity (high/normal) | (1, 15) | 7.87 | .013* |
| **Within-subject** | Connectivity to tumor (connected/unconnected) | (1, 15) | 5.97 | .027* |
| **Interaction** | Peritumor activity x Connectivity to tumor | (1, 15) | 11.02 | .005* |
| Model 2: PATNET |  |  |  |  |
| **Between-subject** | PATNET (strong/weak) | (1, 15) | 1.23 | .285 |
| **Within-subject** | Connectivity to tumor (connected/unconnected) | (1, 15) | 3.97 | .065 |
| **Interaction** | PATNET x Connectivity to tumor | (1, 15) | 3.33 | .088 |

Notes We used a type 3 mixed ANOVA. \* $P \leq .05$ . Levene's test indicated unequal variance for Model 1. However, given the near-equal group sizes for the between-subject factor ( $n = 8$ and $n = 9$ ), the ANOVA is considered robust to this violation. For Model 2 we observed slight non-normality of residuals.

We then checked whether distance to the tumor could be driving deviant activity in tumor-connected distant regions (see supplementary materials for detailed methods). Unsurprisingly, tumor-connected regions were spatially somewhat closer to the tumor (minimum Euclidean distance of median 37.85 mm) than unconnected regions (minimum distance of median 81.28 mm). A linear mixed model (restricted maximum likelihood (REML)) with patients as random effects showed that proximity to the tumor did relate to activity deviance in connected regions as per the standardized coefficient (β *=* −0.012, *SE =* 0.002, *z* = −4.94, *P* < .001): for regions 40mm further from the tumor (i.e. the difference between connected and unconnected regions), the predicted difference in activity deviance was < 0.5 z-score units. These analyses underline that although the connected regions were generally closer to the tumor than all other, unconnected regions (likely based on metabolic constraints and general rules of axonal connectivity favoring shorter connections), proximity to the tumor only explained some of the deviant activity that we observed in connected regions, particularly as most of these regions were still centimeters away from the tumor.

We then explored how tumor volume (corrected for head size), age at diagnosis, sex and having epilepsy might relate to activity deviance in tumor-connected regions, but found no significant associations (Table S3). Specifically, the finding that tumor volume did not relate to activity in tumor-connected regions (*rho*(15) = 0.37, *P* = .149) indicates that this distant hyperactivity was not merely driven by large tumors, but that the structural connection to the tumor itself was most relevant.

Lastly, Cox proportional hazards analyses revealed non-significant hazards ratios in terms of PFS and OS for deviance of activity in distant tumor-connected regions (Table S4).

## Discussion

We here addressed a critical gap in understanding how glioblastomas’ structural embedding into the whole brain shapes peritumor and distant hyperactivity, as well as clinical performance status. First, we found that glioblastomas preferentially occur in locations with intrinsically high structural connectivity as measured in healthy people. Second, structural tumor embedding also explained a large proportion of variance in peritumor hyperactivity, with more embedded tumors showing greater hyperactivity. Moreover, the combination of peritumor hyperactivity and structural connectivity to the tumor explained hyperactivity in distant regions, beyond proximity to the tumor. Finally, stronger tumor embedding based on patient-specific tractography was associated with worse clinical performance.

It is known that gliomas preferentially arise in certain brain regions, namely those with intrinsically higher brain activity, greater functional connectivity and particular genetic signatures.^21,22,46,47^ Here, we showed that glioblastomas also grow more often in locations marked by a higher density of structural streamlines in the healthy brain. These highly structurally embedded regions may thus present a favorable environment for glioblastoma from which to grow and invade the brain, potentially via the emergence of neuron-glioma interactions and the migration of glioma cells via white matter bundles.^24,25,48^

Further focusing on this structural embedding, we then show that such embedding can be established by using a normative tractogram,^24,25^ or by performing tractography on patients’ own diffusion MRI data. Interestingly, we found that normative embedding correlated strongly with patient-specific embedding, negating complete remodeling of tumor connectivity around the glioblastoma. Other literature has reported on significantly lower strength of structural connections in glioblastoma patients versus healthy controls^49^, but our more topologically oriented analysis reveals that the overall connectivity of the peritumor area is largely preserved.

Regarding the clinical relevance of structural embedding, this seems more important for patient-specific tumor embedding as compared to normative embedding. Recent studies showed the clinical relevance of L-TDI for overall survival in glioblastoma,^24,25^ but we could not replicate these reported effects (hazards ratio of ∼1.07 for L-TDI) in our smaller sample. Interestingly, PATNET did relate to the functional performance status of patients, with more strongly embedded tumors relating to lower KPS, potentially reflecting more global brain involvement in such patients. Interestingly, only patient-specific, but not normative structural embedding related to KPS, potentially indicating the added clinical value of patient-specific data. We also found that older individuals tended to have lower PATNET. Since age did not relate to normative tumor embedding, this likely reflects either the known general decline in structural connectivity with age,^50^ or an interaction between healthy aging and tumor growth. Future studies should clarify whether the clinical relevance of patient-specific tumor embedding varies by age.

Turning to the relationship between tumor embedding and peritumor hyperactivity, we confirmed our hypothesis that stronger embedding relates to greater hyperactivity. Critically, healthy activity at individual tumor sites showed no relationship with patients’ peritumor hyperactivity, ruling out a simpler explanation: that hyperactivity merely reflects *where* the tumor is located rather than *how* it is structurally integrated into surrounding tissue. In light of the neuron-glioma interactions that have been observed at the cellular level,^8–10^ our findings support the idea that more highly embedded glioblastomas receive more neuronal input, even from regions that are located further away than just the surrounding tissue.^17,18^

Beyond hyperactivity, structural embedding of cortical regions generally also tracks with functional connectivity, metabolism, and vascularization, all suggesting that more embedded tumors may engage in richer, more varied crosstalk with the surrounding brain tissue.^51–54^

Finally, we shed light on why distant cortical regions are hyperactive in some glioblastoma patients.^9,11^ As hypothesized, distant hyperactivity appears largely confined to patients who already show peritumor hyperactivity. Within those patients, predominantly tumor-connected regions are affected, while unconnected regions are not. These results therefore support the idea that peritumor hyperactivity ‘advances’ via structural connections to more distant brain regions, although our cross-sectional design precludes definitive causal conclusions. Tumor-connected regions may become hyperexcitable through neurogliomal signaling.^8,9,17^ or simply because both normal brain activity^55–57^ and pathological brain activity^58–60^ spread via structural connections. To distinguish between these explanations, non-invasive transcranial magnetic stimulation could test whether low frequency, thus hypothesized inhibitory, stimulation of tumor-connected but not unconnected regions dampens peritumor hyperactivity. Conversely, longitudinal tracking of brain activity and structural connectivity in patients who do not undergo resection could further clarify the temporal sequence: does (increasing) hyperactivity in tumor-connected regions precede (increasing) peritumor hyperactivity, or vice versa? Another interesting future perspective is the investigation of oncological treatments that impact structural connectivity, such as tumor resection and radiotherapy, and their effects on peritumor and distant hyperactivity.

A few limitations of this study should be mentioned. Firstly, we only had a sample of 17 patients available for the PATNET analyses, although it was as homogenous as possible in terms of tumor type and phase of the disease. Also, we included mostly well-functioning patients who were able to provide informed consent and undergo research measurements, potentially leading to a selection bias. It would be important for future research to also capture patients with more symptoms and worse clinical condition. Finally, we used different tumor components for different analyses in this study in order to minimize measurement artifacts per modality, but this choice could induce some divergence between findings, for instance between peritumor hyperactivity and structural embedding.

## Conclusion

We have shown that glioblastoma structural embedding explains hyperactivity both around the tumor and in distant regions, and that this embedding relates to patients’ functional status. These findings bring us closer to understanding how glioblastoma’s integration into the broader brain network shapes its function and patients’ clinical outcome.

## Data availability

Data can be made available in processed and anonymized form under reasonable request. All scripts used for this manuscript can be found on our groups’ public github repository: https://github.com/multinetlab-amsterdam/projects/glioblastoma_embedding _2026

## Competing interests

Mona LM Zimmermann: None to declare

Eva Koderman: One of the inventors on an issued patent related to the application of the canonical correlation analysis to electroencephalography signals.

Marike R van Lingen: Grant MEGIN – Payment to institution for research project on glioma

Lucas C Breedt: None to declare

Dorien Maas: None to declare Niels Verburg: None to declare

Philip de Witt Hamer: None to declare

Arjan Hillebrand: Grant MEGIN – Payment to institution for research project on glioma

Linda Douw: Grant MEGIN – Payment to institution for research project on glioma

## Supplementary material

Supplementary material is available at *Brain* online.

## References

1. Wen YP, Weller M, Lee QE, et al. Glioblastoma in adults: A Society for Neuro-Oncology (SNO) and European Society of Neuro-Oncology (EANO) consensus review on current management and future directions. Neuro-Oncology. 2025;27(11):2751–2788.

2. Kirkman AM, Hunn MHB, Thomas CSM, Tolmie KA. Influences on cognitive outcomes in adult patients with gliomas: A systematic review. Front Oncol. 2022;12

3. Sokolov E, Dietrich J, Cole JA. The complexities underlying epilepsy in people with glioblastoma. The Lancet Neurology. 2023;22(6):505–516.

4. Maas AD, Douw L. Multiscale network neuroscience in neuro-oncology: How tumors, brain networks, and behavior connect across scales. Neuro-Oncology Practice. 2023;10(6):506–517.

5. Douw L, Reijneveld CJ, Mandal SA. Multiscale network perspectives on glioma: from tumour biology to symptoms, survival and treatment. Nature Reviews Neurology. 2026;22(2):73–89.

6. Herr S, Oten S, Bernstock DJ, Rolston DJ, Hervey-Jumper LS. Leveraging Neuroscience Imaging and Electrophysiological Assessments to Decode Neuron-Glioma Interactions. Neuro-Oncology Advances. 2026;

7. Monje M. The neuroscience of brain cancers. Neuron. 2025;113(17):2734–2739.

8. Venkatesh HS, Morishita W, Geraghty AC, et al. Electrical and synaptic integration of glioma into neural circuits. Nature. 2019;573(7775):539–545.

9. Krishna S, Choudhury A, Keough MB, et al. Glioblastoma remodelling of human neural circuits decreases survival. Nature. 2023;617(7961):599–607.

10. Venkataramani V, Yang Y, Schubert MC, et al. Glioblastoma hijacks neuronal mechanisms for brain invasion. Cell. 2022;185(16):2899–2917.e31.

11. Numan T, Kulik DS, Moraal B, et al. Non-invasively measured brain activity and radiological progression in diffuse glioma. Scientific Reports. 2021;11(1)

12. Derks J, Wesseling P, Carbo EWS, et al. Oscillatory brain activity associates with neuroligin-3 expression and predicts progression free survival in patients with diffuse glioma. J Neurooncol. 2018;140(2):403–412.

13. Zimmermann MLM, Breedt CL, Centeno ZGE, et al. The relationship between pathological brain activity and functional network connectivity in glioma patients. J Neurooncol. 2024;166(3):523–533.

14. Belgers V, Numan T, Kulik SD, et al. Postoperative oscillatory brain activity as an add-on prognostic marker in diffuse glioma. J Neurooncol. 2020;147 (1):49–58.

15. Sibih EY, Dada OA, Cunningham EE, et al. Aperiodic neural dynamics define a novel signature of glioma-induced excitation-inhibition dysregulation. bioXiv2025.[Preprint]. doi: 10.1101/2025.05.23.655626.

16. Aabedi AA, Lipkin B, Kaur J, et al. Functional alterations in cortical processing of speech in glioma-infiltrated cortex. Proceedings of the National Academy of Sciences. 2021;118(46):e2108959118.

17. Huang-Hobbs E, Cheng Y-T, Ko Y, et al. Remote neuronal activity drives glioma progression through SEMA4F. Nature. 2023;619(7971):844–850.

18. Sun Y, Wang X, Zhang YD, et al. Brain-wide neuronal circuit connectome of human glioblastoma. Nature. 2025;641(8061):222–231.

19. Derks J, Kulik SD, Numan T, et al. Understanding Global Brain Network Alterations in Glioma Patients. Brain Connectivity. 2021;11(10):865–874.

20. Bosma I, Reijneveld JC, Klein M, et al. Disturbed functional brain networks and neurocognitive function in low-grade glioma patients: a graph theoretical analysis of resting-state MEG. Nonlinear Biomed Phys. 2009;3(1):9.

21. Romero-Garcia R, Mandal AS, Bethlehem RAI, Crespo-Facorro B, Hart MG, Suckling J. Transcriptomic and connectomic correlates of differential spatial patterning among gliomas. Brain. 2023;146(3):1200–1211.

22. Mandal SA, Romero-Garcia R, Hart GM, Suckling J. Genetic, cellular, and connectomic characterization of the brain regions commonly plagued by glioma. Brain. 2020;143(11):3294–3307.

23. Salvalaggio A, Pini L, Bertoldo A, Corbetta M. Glioblastoma and brain connectivity: the need for a paradigm shift. The Lancet Neurology. 2024;23(7):740–748.

24. Salvalaggio A, Pini L, Gaiola M, et al. White Matter Tract Density Index Prediction Model of Overall Survival in Glioblastoma. JAMA Neurology. 2023;80(11):1222–1231.

25. Falcó-Roget J, Basile AG, Janus A, et al. A non-local diffusion magnetic resonance imaging tract density biomarker to stratify, predict, and interpret survival rates in human glioblastoma. Neuro-Oncology. 2026;28(2):564–579.

26. Louis DN, Perry A, Wesseling P, et al. The 2021 WHO Classification of Tumors of the Central Nervous System: a summary. Neuro-Oncology. 2021;23(8):1231–1251.

27. Breedt CL, Santos NAF, Hillebrand A, et al. Multimodal multilayer network centrality relates to executive functioning. Network Neuroscience. 2023;7(1):299–321.

28. Bakas S, Akbari H, Sotiras A, et al. Advancing The Cancer Genome Atlas glioma MRI collections with expert segmentation labels and radiomic features. Scientific Data. 2017;4(1):170117.

29. Clark K, Vendt B, Smith K, et al. The Cancer Imaging Archive (TCIA): Maintaining and Operating a Public Information Repository. Journal of Digital Imaging. 2013;26(6):1045–1057.

30. Pemberton GH, Wu J, Kommers I, et al. Multi-class glioma segmentation on real-world data with missing MRI sequences: comparison of three deep learning algorithms. Scientific Reports. 2023;13(1)18911.

31. Fan L, Li H, Zhuo J, et al. The Human Brainnetome Atlas: A New Brain Atlas Based on Connectional Architecture. Cerebral Cortex. 2016;26(8):3508–3526.

32. Jenkinson M, Smith S. A global optimisation method for robust affine registration of brain images. Medical Image Analysis. 2001;5(2):143–156.

33. Jenkinson M. Improved Optimization for the Robust and Accurate Linear Registration and Motion Correction of Brain Images. NeuroImage. 2002;17(2):825–841.

34. Elias BJG, Germann J, Joel ES, et al. A large normative connectome for exploring the tractographic correlates of focal brain interventions. Scientific Data. 2024;11353.

35. Billot B, Magdamo C, Cheng Y, Arnold ES, Das S, Iglesias EJ. Robust machine learning segmentation for large-scale analysis of heterogeneous clinical brain MRI datasets. Proceedings of the National Academy of Sciences. 2023;120(9)

36. Billot B, Greve ND, Puonti O, et al. SynthSeg: Segmentation of brain MRI scans of any contrast and resolution without retraining. Medical Image Analysis. 2023;86:102789.

37. Iglesias EJ, Billot B, Balbastre Y, et al. SynthSR: A public AI tool to turn heterogeneous clinical brain scans into high-resolution T1-weighted images for 3D morphometry. Science Advances. 2023;9(5)

38. Gopinath K, Greve, D.N., Das, S., Arnold, S., Magdamo, C., Iglesias, J.E. Cortical analysis of heterogeneous clinical brain MRI scans for large-scale neuroimaging studies. presented at: Medical Image Computing and Computer Assisted Intervention – MICCAI 2023 MICCAI 2023 Lecture Notes in Computer Science; 2023; https://arxiv.org/abs/2305.01827

39. Smith ER, Tournier J-D, Calamante F, Connelly A. SIFT2: Enabling dense quantitative assessment of brain white matter connectivity using streamlines tractography. NeuroImage. 2015;119:338–351.

40. Klink VN, Rosmalen VF, Nenonen J, et al. Automatic detection and visualisation of MEG ripple oscillations in epilepsy. NeuroImage: Clinical. 2017;15:689–701.

41. Taulu S, Simola J. Spatiotemporal signal space separation method for rejecting nearby interference in MEG measurements. Physics in Medicine and Biology. 2006;51(7):1759–1768.

42. Hillebrand A, Tewarie P, Dellen VE, et al. Direction of information flow in large-scale resting-state networks is frequency-dependent. Proceedings of the National Academy of Sciences. 2016;113(14):3867–3872.

43. Hillebrand A, Barnes GR, Bosboom JL, Berendse HW, Stam CJ. Frequency-dependent functional connectivity within resting-state networks: An atlas-based MEG beamformer solution. NeuroImage. 2012;59(4):3909–3921.

44. Manning JR, Jacobs J, Fried I, Kahana MJ. Broadband Shifts in Local Field Potential Power Spectra Are Correlated with Single-Neuron Spiking in Humans. Journal of Neuroscience. 2009;29(43):13613–13620.

45. Zimmermann MLM, Ulrich C, Breedt CL, et al. The relationship between deviant brain activity and executive functioning in glioma patients. Neuro-Oncology Advances. 2025;7(1)

46. Numan T, Breedt CL, Maciel CPADB, et al. Regional healthy brain activity, glioma occurrence and symptomatology. Brain. 2022;145(10):3654–3665. 0

47. Douw L, Breedt CL, Zimmermann MLM. Cancer meets neuroscience: the association between glioma occurrence and intrinsic brain features. Brain. 2023;146(3):803–805.

48. Scherer. Structural development in Gliomas. The American Journal of Cancer. 1938;34(3):333–351.

49. Wei Y, Li C, Cui Z, et al. Structural connectome quantifies tumour invasion and predicts survival in glioblastoma patients. Brain. 2023;146(4):1714–1727.

50. Zhao T, Cao M, Niu H, et al. Age[related changes in the topological organization of the white matter structural connectome across the human lifespan. Human Brain Mapping. 2015;36(10):3777–3792.

51. Liang X, Zou Q, He Y, Yang Y. Coupling of functional connectivity and regional cerebral blood flow reveals a physiological basis for network hubs of the human brain. Proceedings of the National Academy of Sciences. 2013;110(5):1929–1934.

52. Aslan S, Huang H, Uh J, et al. White matter cerebral blood flow is inversely correlated with structural and functional connectivity in the human brain. NeuroImage. 2011;56(3):1145–1153.

53. Várkuti B, Cavusoglu M, Kullik A, et al. Quantifying the Link between Anatomical Connectivity, Gray Matter Volume and Regional Cerebral Blood Flow: An Integrative MRI Study. PLoS ONE. 2011;6(4):e14801.

54. Ji X, Ferreira T, Friedman B, et al. Brain microvasculature has a common topology with local differences in geometry that match metabolic load. Neuron. 2021;109(7):1168–1187.e13.

55. Lynn WC, Bassett SD. The physics of brain network structure, function and control. Nature Reviews Physics. 2019;1(5):318–332.

56. Seguin C, Sporns O, Zalesky A. Brain network communication: concepts, models and applications. Nat Rev Neurosci. 2023;24(9):557–574.

57. Hahn G, Ponce-Alvarez A, Deco G, Aertsen A, Kumar A. Portraits of communication in neuronal networks. Nat Rev Neurosci. 2019;20(2):117–127.

58. Azeem A, Abdallah C, Ellenrieder VN, Kosseifi EC, Frauscher B, Gotman J. Explaining slow seizure propagation with white matter tractography. Brain. 2024;147(10):3458–3470.

59. Gleichgerrcht E, Greenblatt SA, Kellermann ST, et al. Patterns of seizure spread in temporal lobe epilepsy are associated with distinct white matter tracts. Epilepsy Research. 2021;171:106571.

60. Boddeti U, Farooque P, Mcgrath H, et al. Identifying the epileptic network by linking interictal functional and structural connectivity. Scientific Reports. 2025;15(1)9106.

