## Supplementary materials for "Structural network embedding governs peritumor and distant pathological brain activity in glioblastoma"

Author affiliations:

De Boelelaan 1108, 1081 HZ Amsterdam

**Running title**: Glioblastoma embedding governs hyperactivity

| Table S1 Overview of earlier studies using an overlapping patient cohort | |
| --- | --- |
| **Authors (Year)** | **Title** |
| Douw et al. (2010)^1^ | Epilepsy is related to theta band brain connectivity and network topology in brain tumor patients |
| van Dellen et al. (2012)^2^ | MEG Network Differences between Low- and High-Grade Glioma Related to Epilepsy and Cognition |
| van Dellen et al. (2012)^3^ | Connectivity in MEG resting-state networks increases after resective surgery for low-grade glioma and correlates with improved cognitive performance. |
| Carbo et al. (2017)^4^ | Dynamic hub load predicts cognitive decline after resective neurosurgery |
| Derks et al. (2018)^5^ | Oscillatory brain activity associates with neuroligin-3 expression and predicts progression free survival in patients with diffuse glioma. |
| Derks et al. (2019)^6^ | Understanding cognitive functioning in glioma patients: The relevance of IDH-mutation status and functional connectivity |
| Belgers et al. (2020)^7^ | Postoperative oscillatory brain activity as an add-on prognostic marker in diffuse glioma. |
| Numan et al. (2021)^8^ | Non-invasively measured brain activity and radiological progression in diffuse glioma |
| Derks et al. (2021)^9^ | Understanding Global Brain Network Alterations in Glioma Patients |
| van Lingen et al. (2023)^10^ | The longitudinal relation between executive functioning and multilayer network topology in glioma patients |
| Zimmermann et al. (2024)^11^ | The relationship between pathological brain activity and functional network connectivity in glioma patients |
| Zimmermann et al. (2025)^12^ | The relationship between Between Deviant Brain Activity and Executive Functioning in Glioma Patients |
| *Note.* Table adapted from Röttgering et al. (2024)^13^ and Zimmermann et al. (2025)^11^ | |

| Table S2 TCGA patient characteristics |  |
| --- | --- |
|  | **TCGA glioblastoma patients**  **(N = 102)** |
| Mean age in years (SD) | 58 (14) |
| Number of males \| females \| unknown | 63 \| 38 \| 1 |
| Glioma lateralization left \| right \| bilateral | 63 \| 38 \| 1 |
| Median tumor volume in cm^3^ in MNI space (Q1-Q3) | 53 (26-84) |
| Number of patients with KPS < 90 \| 90-100 \| unknown | 72 \|17 \| 13 |
| Epilepsy (yes/no) | NA |
| SD = standard deviation. Q1-Q3 = quartile 1 -3. KPS = Karnofsky performance status. NA = not available | |

| Table S3 Covariate exploration | | | | | | |
| --- | --- | --- | --- | --- | --- | --- |
|  | Variable | Mean | Standard Deviation | *df* | *Statistical value* | *P*-value |
| Mean BBP_dev_ in tumor regions | Sex (male/female) | 1.56 / 0.67 | 1.65 /1.06 | 27 | *U* = 88 | .328 |
|  | Epilepsy (yes/no) | 1.13 / 1.77 | 1.55 / 1.61 | 27 | *U* = 124 | .276 |
|  | Age (continuous) | - | - | *27* | *rho = -0.01* | .964 |
|  | Tumor volume (continuous) | - | - | *27* | *rho = 0.39* | .039* |
| Mean BBP_dev_ in tumor-connected regions | Sex (male/female) | 0.44 / -0.28 | 1.08 / 0.31 | 15 | *t* = 1.29 | .218 |
|  | Epilepsy (yes/no) | 0.35 / 0.13 | 1.17 / 0.62 | 15 | *U =* 33 | 1.00 |
|  | Age (continuous) | - | - | 15 | *rho = -.28* | .275 |
|  | Tumor volume (continuous) |  |  | 15 | *rho = 0.37* | .149 |
| PATNET | Sex (male/female) | 44.90 / 52.00 | 18.34 / 24.48 | 15 | *t* = -0.63 | .539 |
|  | Epilepsy (yes/no) | 49.63 / 41.00 | 21.77 / 13.83 | 15 | *t* = -0.87 | .396 |
|  | Age (continuous) | - | - | 15 | *rho = -.48* | .050* |
|  | Tumor volume (continuous) | - | - | 15 | *rho=.52* | .032* |
| L-TDI | Sex (male/female) | 66.12 / 45.61 | 29.73 / 27.17 | 27 | *t* = 1.53 | .138 |
|  | Epilepsy (yes/no) | 56.47 / 70.71 | 30.73 / 27.76 | 27 | *t* = 1.25 | .221 |
|  | Age (continuous) | - | - | 27 | *rho = -.08* | .668 |
|  | Tumor volume (continuous) | - | - | 27 | *rho = .86* | <.001** |
| *Notes.* If assumption of normality was violated, Mann-Whitney U tests were used.* P ≤ .05, P < .001**, BBP_dev_ = Broadband power deviance. | | | | | | |

| Table S4 Cox proportional Hazards analyses | | | |
| --- | --- | --- | --- |
| Dependent variable (univariate models) | Predictor | HR [95% CI] | *P*-value |
| PFS | L-TDI | 1.00 [0.99, 1.02] | 0.674 |
|  | Mean BBP_dev_ in tumor connected regions | 1.34 [0.83, 2.15] | 0.227 |
|  | PATNET | 1.01 [0.99, 1.04] | 0.355 |
| OS | L-TDI | 1.01 [0.99, 1.02] | 0.386 |
|  | Mean BBP_dev_ in tumor- connected regions | 1.43 [0.92, 2.24] | 0.111 |
|  | PATNET | 1.02 [0.99, 1.04] | 0.199 |
| *Notes* Models were run separately per predictor. The proportional hazards assumption was violated for predictor L-TDI in both models, suggesting a time-varying effect. | | | |

MNI space used in all analyses

For all analyses where we mention ‘MNI space’ in the main manuscript body, we specifically mean MNI ICBM 2009b NLIN Asymmetric space.

Tumor mask procedure

To define tumor regions based on subject’s thresholded and binarized tumor masks in native space we first skullstripped and segmented the patient’s brain using synthstrip and synthseg from the Free Surfer recon-all-clinical pipeline.^14-17^ For most patients (except for three) synthseg segmented the brain in a resampled space so we used FSL FLIRT to register the segmentation to native space using nearest neighbor interpolation^18,19^. We then thresholded and binarized the output of synthseg to only obtain the gray matter ribbon from the segmentation. Subsequently, we registered the BNA atlas in MNI152 linear space to the subjects’ native space, skull-stripped image. To do so, we first co-registered the subject’s native space, skullstripped image to the MNI brain template to obtain the transformation matrix from native to MNI space using FSL FLIRT.^18,19^ We then used the inverse of the transformation matrix to register the BNA mask from MNI space to the native space with FSL FLIRT, using nearest-neighbor interpolation to preserve the correct labels of the BNA atlas.^18,19^ Then, using fslmaths we multiplied the binarized gray matter ribbon obtained earlier with the BNA atlas to extract the patient-specific, gray matter areas of the BNA atlas regions.^20-22^ Next, we extracted the volume of each of the 210 gray-matter, cortical regions of the BNA atlas using fslstats. As a last step, we again used fslmaths to multiply the binarized tumor mask with each of the 210 BNA regions to define their overlap and extracted the volume of this overlap using fslstats. In further analyses we used the volume of each BNA region and the volume of the tumor overlap with that region, to calculate the percentage of a region that was covered by the tumor ((volume overlap/volume region)*100). We defined regions as tumoral if 20% or more of their volume was covered by tumor. We used a cut-off of 20% based on data explorations on the number subjects that had a tumor in the different regions striving for a balance between having a few regions with a high percentage of subjects with a tumor there and most regions having only a few subjects with a tumor. A cut-off of 20%, provided a good variation in the data (i.e. having enough tumor regions, that show large variation in the number of subjects who show a tumor there). Throughout all steps of this procedure we visually quality controlled the registrations and extractions using fsleyes (<https://zenodo.org/records/18990371>).

Distance analysis of tumor-connected regions

To investigate whether proximity to the tumor, instead of only tumor connectivity, might be driving our findings we extracted the coordinates of the centroids of the 210 cortical BNA regions in MNI152 linear space and calculated the pairwise Euclidean distances between the coordinates of the centroids of all regions. Then, for every patient we extracted the distances of the BNA regions defined as tumor regions (based on the >=20% overlap with the tumor mask) to all non-tumor BNA regions. Finally, for every non-tumor region, we extracted the minimum distance between that region to any of the tumor regions.

References

1.	Douw L, van Dellen E, de Groot M, et al. Epilepsy is related to theta band brain connectivity and network topology in brain tumor patients. *BMC Neurosci*. 2010;11(1):103.

2.	van Dellen E, Douw L, Hillebrand A, et al. MEG Network Differences between Low- and High-Grade Glioma Related to Epilepsy and Cognition. *PLoS ONE*. 2012;7(11):e50122.

3.	van Dellen E, de Witt Hamer PC, Douw L, et al. Connectivity in MEG resting-state networks increases after resective surgery for low-grade glioma and correlates with improved cognitive performance. *NeuroImage: Clinical*. 2012;2:1–7.

4.	Carbo EWS, Hillebrand A, van Dellen E, et al. Dynamic hub load predicts cognitive decline after resective neurosurgery. *Scientific Reports*. 2017;7(1):42117.

5.	Derks J, Wesseling P, Carbo EWS, et al. Oscillatory brain activity associates with neuroligin-3 expression and predicts progression free survival in patients with diffuse glioma. *J Neurooncol*. 2018;140(2):403–412.

6.	Derks J, Kulik S, Wesseling P, et al. Understanding cognitive functioning in glioma patients: The relevance of IDH‐mutation status and functional connectivity. *Brain Behav*. 2019;9(4):e01204.

7.	Belgers V, Numan T, Kulik SD, et al. Postoperative oscillatory brain activity as an add-on prognostic marker in diffuse glioma. *J Neurooncol*. 2020;147(1):49–58.

8.	Numan T. Non-invasively measured brain activity and radiological progression in diffuse glioma. *Scientific Reports*. 2021 2021;11:10. 18990.

9.	Derks J, Kulik SD, Numan T, et al. Understanding Global Brain Network Alterations in Glioma Patients. *Brain Connectivity*. 2021;11(10):865–874.

10.	Lingen VRM, Breedt CL, Geurts JGJ, et al. The longitudinal relation between executive functioning and multilayer network topology in glioma patients. *Brain Imaging and Behavior*. 2023;17(4):425–435.

11.	Zimmermann MLM, Breedt CL, Centeno ZGE, et al. The relationship between pathological brain activity and functional network connectivity in glioma patients. *J Neurooncol*. 2024;166(3):523–533.

12.	Zimmermann MLM, Ulrich C, Breedt CL, et al. The relationship between deviant brain activity and executive functioning in glioma patients. *Neuro-Oncology Advances*. 2025;7(1)

13.	Röttgering JG, Varkevisser TMCK, Gorter M, et al. Symptom networks in glioma patients: understanding the multidimensionality of symptoms and quality of life. *J Cancer Surviv*. 2024;18(3):1032–1041.

14.	Billot B, Magdamo C, Cheng Y, Arnold ES, Das S, Iglesias EJ. Robust machine learning segmentation for large-scale analysis of heterogeneous clinical brain MRI datasets. *Proceedings of the National Academy of Sciences*. 2023;120(9)

15.	Billot B, Greve ND, Puonti O, et al. SynthSeg: Segmentation of brain MRI scans of any contrast and resolution without retraining. *Medical Image Analysis*. 2023;86:102789.

16.	Iglesias EJ, Billot B, Balbastre Y, et al. SynthSR: A public AI tool to turn heterogeneous clinical brain scans into high-resolution T1-weighted images for 3D morphometry. *Science Advances*. 2023;9(5)

17.	Gopinath K, Greve, D.N., Das, S., Arnold, S., Magdamo, C., Iglesias, J.E. Cortical analysis of heterogeneous clinical brain MRI scans for large-scale neuroimaging studies. presented at: Medical Image Computing and Computer Assisted Intervention – MICCAI 2023 MICCAI 2023 Lecture Notes in Computer Science; 2023; <https://arxiv.org/abs/2305.01827>

18.	Jenkinson M, Smith S. A global optimisation method for robust affine registration of brain images. *Medical Image Analysis*. 2001;5(2):143–156.

19.	Jenkinson M. Improved Optimization for the Robust and Accurate Linear Registration and Motion Correction of Brain Images. *NeuroImage*. 2002;17(2):825–841.

20.	Woolrich WM, Jbabdi S, Patenaude B, et al. Bayesian analysis of neuroimaging data in FSL. *NeuroImage*. 2009;45(1):S173–S186.

21.	Smith MS, Jenkinson M, Woolrich WM, et al. Advances in functional and structural MR image analysis and implementation as FSL. *NeuroImage*. 2004;23:S208–S219.

22.	Jenkinson M, Beckmann FC, Behrens EJT, Woolrich WM, Smith MS. FSL. *NeuroImage*. 2012;62(2):782–790.
